# Graph-Augmented Retrieval for Digital Evidence-Based Medical Synthesis: A Proof-of-Concept Study on Topology-Aware Mechanistic Narrative Generation

**DOI:** 10.64898/2026.02.18.26346545

**Authors:** Filippo Buscemi, Primo Buscemi

## Abstract

**Background:** Retrieval-augmented generation (RAG) frameworks such as RAPID [1] have demonstrated that staged planning and retrieval grounding improve long-form text generation. However, most implementations remain similarity-driven and open-domain, lacking the epistemic safeguards required for biomedical synthesis, where mechanistic completeness, temporal governance, traceability, and explicit gap classification are essential.

**Objective:** To develop and evaluate a topology-aware, graph-augmented retrieval framework for structured biomedical narrative synthesis, and to position it as a domain-constrained evolution of staged RAG aligned with structural principles of digital evidence-based medicine (dEBM).

**Methods:** We implemented a two-layer architecture operating on a closed, version-controlled corpus of 11,861 peer-reviewed text chunks on iron deficiency. A metadata-constrained vector retriever (RAG01) was extended with a Graph-RAG (RAG02) overlay (RAG02) constructed from chunk-level entity extraction and weighted co-occurrence networks (30 nodes; 118 directed edges). Topic planning was organized through predefined mechanistic axes functioning as structured hypothesis probes. Retrieval was performed under identical deterministic constraints (top-*k* = 5; cosine threshold = 0.50; publication year ≥ 2023), and graph diagnostics—including local connectivity, induced subgraph density, modular overlap, and multi-hop stability—were used to distinguish retrieval insufficiency from corpus-level evidentiary scarcity.

**Results:** In a case study of obesity-associated iron deficiency, the entity network exhibited a centralized regulatory topology with hepcidin as a high-connectivity hub. Axis-based retrieval combined with graph auditing consistently reinforced an inflammation-mediated hepcidin pathway linking obesity to iron deficiency, while alternative mechanisms lacked stable multi-hop embedding. Compared with vector-only retrieval, graph augmentation preserved semantic alignment and increased mean cosine similarity from 0.673 to 0.694 while reducing similarity dispersion (SD 0.056 to 0.035) under identical constraints. Graph activity ratio was 1.00 in the temporally filtered corpus.

**Conclusions:** By integrating mechanistic axis decomposition, topology-aware auditing, causal scaffolding, and expert-driven iterative refinement, the proposed framework implements selected structural constraints inspired by evidence-based medicine within a controlled digital synthesis environment. The approach advances retrieval-augmented generation beyond similarity-based summarization toward a reproducible model of topology-aware biomedical evidence interrogation with implications for AI-assisted systematic reviews.

## 1 Introduction

The generation of long, knowledge-intensive documents remains a central challenge for large language models (LLMs). While recent advances have substantially improved short-form reasoning and summarization, the production of coherent, factually grounded, and structurally consistent long-form scientific texts continues to expose limitations related to hallucination, topic drift, and epistemic instability, issues that have recently motivated dedicated faithfulness-detection frameworks. [2] In high-stakes domains such as biomedicine, these limitations are not merely technical artifacts but epistemic risks, as narrative amplification of weakly supported mechanisms may distort the state of evidence.

Retrieval-Augmented Generation (RAG) has emerged as a practical strategy to mitigate hallucinations by grounding model outputs in external corpora. However, most RAG implementations are optimized for short-answer question answering or open-domain content generation. Longform biomedical synthesis introduces additional constraints: comprehensive mechanistic coverage, cross-sectional logical coherence, reproducible evidence traceability, and explicit control of unsupported hypotheses. Similarity-based retrieval alone does not guarantee that retrieved concepts are structurally reinforced within the underlying literature, and recent work has shown that redundancy-aware and set-level selection objectives can materially alter downstream generation behavior [3]. Alternative embedding strategies incorporating topic-level structure have been proposed to mitigate such limitations [4].

The RAPID framework demonstrated that staged planning and dependency-aware drafting improve long-form generation coherence. More recently, graph-based RAG architectures have incorporated structured representations to enhance reasoning reliability and evidence governance.[5][6] Related efforts extend retrieval memory into higherorder graph or hypergraph structures to support multi-step reasoning.[7] EBM-oriented adaptations, such as SR-RAG, integrate PICO alignment and evidence hierarchy into retrieval and reranking strategies.[8] These approaches improve population-level alignment and grading consistency, yet mechanistic completeness and structural reinforcement of biological pathways remain comparatively underexplored dimensions of retrieval evaluation.

Biomedical synthesis differs from encyclopedic generation in that completeness is not defined solely by section coverage or document-level relevance, but by systematic interrogation of plausible biological mechanisms.[9] Semantically relevant excerpts may correspond to hypotheses that are sparsely embedded or weakly reinforced within the corpus topology. In such cases, similarity-driven aggregation risks conflating narrative plausibility with corpus-level support.

In this work, we propose a domain-constrained evolution of staged retrieval-augmented long-form generation tailored to biomedical evidence synthesis. We introduce a two-layer architecture: (1) RAG01, a metadata-constrained vector retrieval system operating on a curated and temporally governed biomedical corpus; and (2) Graph-RAG (RAG02), an entity-centric graph overlay enabling multi-hop structural auditing, mechanistic prioritization, and explicit classification of unsupported axes. Unlike PICO-centered adaptations, our framework adopts mechanistic axis decomposition as the primary planning scaffold.Each predefined axis functions as an epistemic probe whose support is evaluated through both similarity retrieval and graph-topological reinforcement, conceptually aligning with early alignment strategies between planning and retrieval [10].

We define topology-aware retrieval as a paradigm in which ranking and mechanism prioritization are influenced not only by semantic similarity but also by the measurable structural embedding of concepts within a corpus-derived knowledge graph. Structural coherence is quantified through connectivity, induced subgraph density, modular overlap, and multi-hop stability diagnostics. Importantly, topology-awareness does not equate structural centrality with biological truth; rather, it introduces an explicit structural validation layer that distinguishes retrieval insufficiency from corpus-level evidentiary scarcity.

The proposed framework is evaluated through a controlled case study on obesity-associated iron deficiency. This domain provides a suitable test scenario for mechanistic discrimination, as multiple biologically plausible pathways have been proposed. Evaluation focuses on comparative structural behavior under identical deterministic retrieval constraints rather than on large-scale benchmark optimization. Specifically, we assess whether graph augmentation preserves semantic alignment while improving structural coherence and axis-level discrimination within a fixed corpus snapshot.

By integrating deterministic planning, temporal governance, graph-theoretic auditing, causal scaffolding, and specialist validation, this work advances retrieval-augmented generation from similarity-based aggregation toward topology-aware biomedical evidence interrogation. The contribution is methodological: we demonstrate how structural constraints can be incorporated into long-form retrieval workflows to improve epistemic transparency and mechanistic discrimination in AI-assisted biomedical synthesis.

## 2 Methods

### 2.1 Study Objective and Conceptual Architecture

The objective of this study was to design and evaluate a domain-specific retrieval–generation framework for structured, evidence-traceable biomedical synthesis on iron deficiency.

The system integrates two retrieval layers operating on the same indexed corpus, conceptually related to hierarchical and multi-pass filtering paradigms in recent RAG architectures [11]:

- **RAG01**: metadata-constrained dense vector retrieval over the full embedding space.
- **Graph-RAG (RAG02)**: an entity-centric graph overlay enabling structural auditing, multi-hop expansion, and mechanistic gap classification.

**Figure 1:**
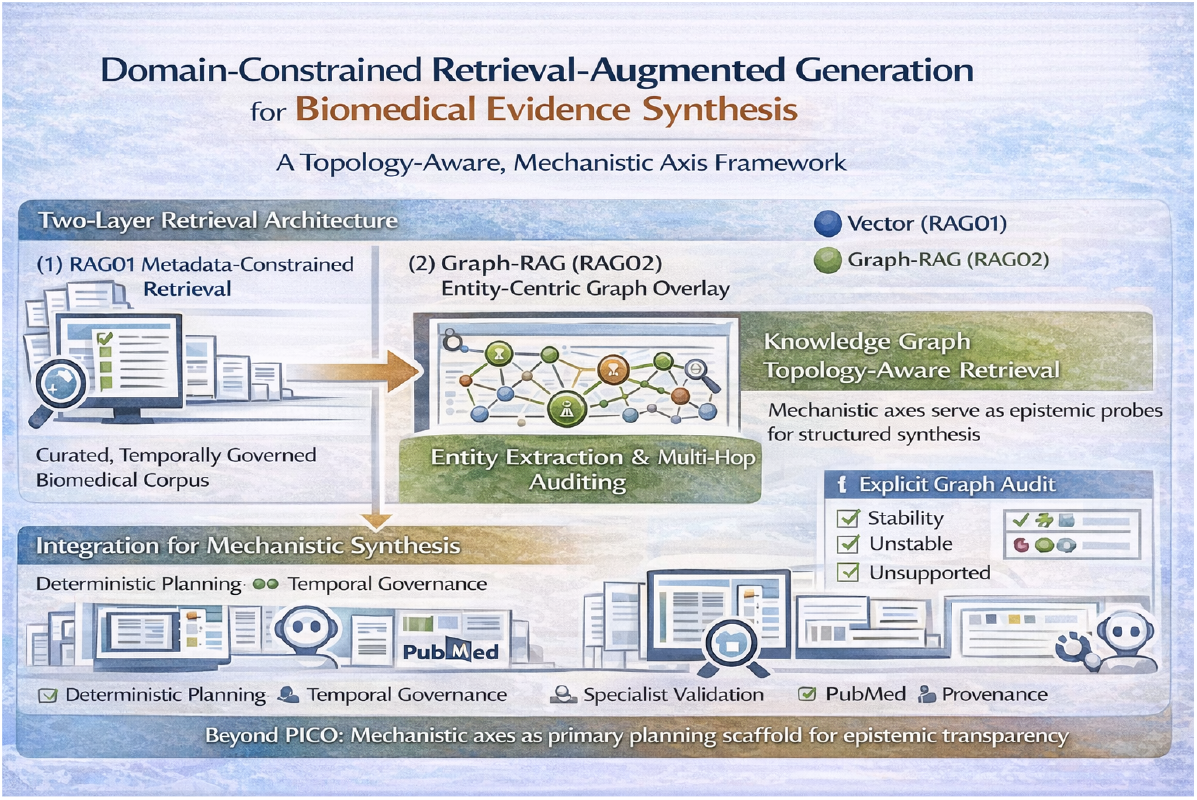
Conceptual overview of the proposed domain-constrained retrieval framework. The architecture integrates two retrieval layers operating on a curated and temporally governed biomedical corpus: (1) RAG01, a metadata-constrained dense vector retriever, and (2) Graph-RAG (RAG02), an entity-centric graph overlay enabling multi-hop structural auditing. Unlike PICO-centered adaptations, planning is organized through predefined mechanistic axes that function as epistemic probes. Axis support is evaluated through the interaction between semantic similarity retrieval and graph-topological reinforcement, introducing an explicit structural validation layer within a closed-corpus environment.

Formally, we define:

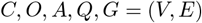

where:

- *C* denotes the indexed biomedical corpus,
- *O* the predefined outline scaffold,
- *A* the set of mechanistic axes,
- *Q* the structured retrieval queries,
- *G* = (*V, E*) the entity graph derived from chunk-level co-occurrence.

#### Formal Hypothesis

Under an identical corpus snapshot and identical retrieval constraints, the graph-augmented configuration (RAG02) was hypothesized to improve structural coherence of mechanistic axes and reinforce embedding around central regulatory nodes compared with vector-only retrieval (RAG01). The objective was structural comparison rather than generative fluency assessment.

### 2.2 Corpus Construction and Indexing Pipeline

#### 2.2.1 Document Parsing and Normalization

Source PDF publications were processed using **GROBID (v0.8.x)** to extract structured TEI XML. Outputs were normalized through a custom TEI-to-JSON pipeline (tei-normalizer v2), which:

- segmented documents into section- and paragraph-level chunks,
- preserved semantic hierarchy,
- standardized metadata fields,
- generated stable `chunk_key` identifiers,
- validated publication year using TEI and Crossref.

Each normalized textual unit constitutes a retrieval chunk.

#### 2.2.2 Corpus Composition

The indexed corpus comprises:

- 11,861 text chunks
- 627 unique peer-reviewed publications
- Publication years spanning 2018–2026

DOI completeness was 100%. Validated publication year metadata was missing in 3.1% of chunks and retained as null without imputation.

Chunk length ranged from 46 to 3,990 characters (mean 1,271).

#### Embedding Model and Vector Space

All embeddings were generated using the `OpenAI text-embedding-3-large` model (3,072 dimensions). Embeddings were L2-normalized prior to storage, ensuring cosine similarity equivalence with inner-product ranking in pgvector.

Vector dimensionality was fixed per corpus snapshot. No mixed-model embeddings or dynamic re-embedding were performed. Similarity ranking was executed entirely within PostgreSQL using pgvector’s cosine operator.

### 2.3 Graph Construction

Entity extraction identified:

- 10,393 entity mentions,
- 4,887 graph-active chunks (41.2% of corpus),
- 30 normalized biomedical entities (nodes),
- 118 directed weighted co-occurrence edges.

Edges represent chunk-level co-occurrence frequency between entity pairs. The graph is fully directional (reciprocity ratio *R* = 0), excluding symmetric inflation.

#### Graph Construction Rules

- Minimum co-occurrence threshold: ≥ 3
- Self-loops excluded
- Directionality preserved
- No runtime pruning during evaluation

Graph structure was treated as a descriptive structural layer and not optimized post hoc.

### 2.4 Retrieval Configurations

#### 2.4.1 RAG01 — Vector Retrieval

Given query embedding *q* ∈ ℝ^*d*^ and chunk embedding *c*_*i*_ ∈ ℝ^*d*^

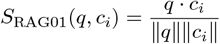

Ranking was performed via descending cosine similarity.

#### 2.4.2 RAG02 — Hybrid Graph-Augmented Retrieval

Graph-aware scoring was defined as [12]:

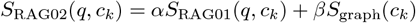

where:

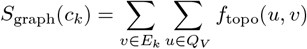

Hybrid weights were fixed a priori:

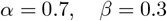

No parameter tuning was performed.

### 2.5 Experimental Protocol

Evaluation was conducted using eight predefined mechanistic queries addressing obesity-associated iron deficiency.

Identical retrieval constraints were applied to both systems:

- top-*k* = 5
- cosine similarity threshold = 0.50
- publication_year_int ≥ 2023
- fixed corpus snapshot (11,861 chunks)

All runs were deterministic. No stochastic sampling, adaptive learning, or dynamic re-ranking was used.

### 2.6 Quantitative Retrieval Evaluation

To compare structural and semantic behavior between RAG01 and RAG02, query-level retrieval outputs were aggregated.

For each query, the top-*k* chunks were recorded and the following metrics computed:

- **Mean Cosine Similarity (MCS)** — average cosine similarity of retrieved chunks.
- **Similarity Dispersion (SD)** — standard deviation of similarity scores.
- **Graph Activity Ratio (GAR)** — proportion of retrieved chunks present in the graph-active subset.

Metrics were aggregated across queries to generate system-level summaries.

The evaluation objective was structural comparison of retrieval behavior rather than statistical hypothesis testing, aligning with interpretable and comparative evaluation paradigms recently proposed for RAG systems [13].

### 2.7 Topological Diagnostics

Structural embedding was assessed using:

#### 1-hop neighborhood

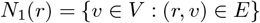

#### Induced subgraph density (directed)

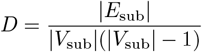

#### Jaccard overlap

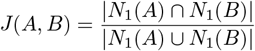

These diagnostics were used to classify mechanistic axes as structurally supported, weakly supported, or unsupported within the corpus snapshot.[8]

### 2.8 Reproducibility and Determinism

All experiments were conducted on a fixed corpus snapshot within a PostgreSQL (pgvector-enabled) environment. Identical queries under identical parameters produce identical retrieval outputs, a design choice aligned with emerging work on robustness and certified stability in neural ranking systems [14]. No external APIs, adaptive re-ranking, or runtime graph mutation were employed.

This design ensures full traceability from retrieved chunk to final narrative synthesis.

## 3. Results

### 3.1 Corpus-Level Graph Architecture

The audited core entity graph comprised 30 normalized biomedical entities connected by 118 directed, weighted co-occurrence edges (reciprocity ratio *R* = 0), excluding symmetric artifact inflation.

Weighted degree analysis identified a centralized iron-regulatory topology. The dominant regulatory core included:

- Ferritin
- Iron deficiency
- Hemoglobin
- Hepcidin
- Erythropoiesis

Hepcidin exhibited high local connectivity within the audited graph, consistent with its established regulatory role in iron metabolism. By contrast, obesity occupied a less central but structurally embedded position within the network.

These findings indicate a topology organized around iron-regulatory hubs, with condition-specific entities positioned within peripheral yet connected modules.

To assess whether this structural organization translated into mechanistic discrimination at the condition level, we conducted axis-based retrieval combined with graph-topological auditing.

### 3.2 Mechanistic Axis Evaluation: Obesity–Iron Deficiency Case Study

Structured axis-based retrieval combined with graph-topological auditing yielded differentiated patterns of support across hypothesized mechanisms.

#### Inflammation–Hepcidin Axis

This axis demonstrated robust structural embedding within the entity network.

High-weight directed edges connected hepcidin with iron deficiency, iron absorption, IL-6, and obesity, forming a stable multi-hop pathway consistent with the sequence:

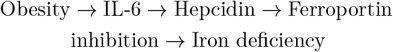

Multi-hop expansion consistently reproduced this regulatory cascade. The convergence of retrieval density, local connectivity, and reproducible multihop reinforcement supports inflammation-mediated hepcidin upregulation as the dominant corpus-level mechanism linking obesity to iron deficiency.

#### Hepcidin-Independent Transporter Mechanisms

Neither 1-hop nor 2-hop expansion demonstrated stable structural embedding between obesity and intestinal transporter nodes. Although semantically relevant excerpts were retrievable under dense similarity alone, graph-level reinforcement was absent.

This axis was therefore classified a s a structural corpus gap rather than a retrieval failure.

#### Marrow Remodeling Axis

Erythropoietic nodes displayed global centrality within the core graph. However, specific linkage between obesity and marrow remodeling pathways was weak and inconsistently reinforced across multihop expansion.

This axis was classified as weakly supported within the current corpus snapshot.

**Figure 2:**
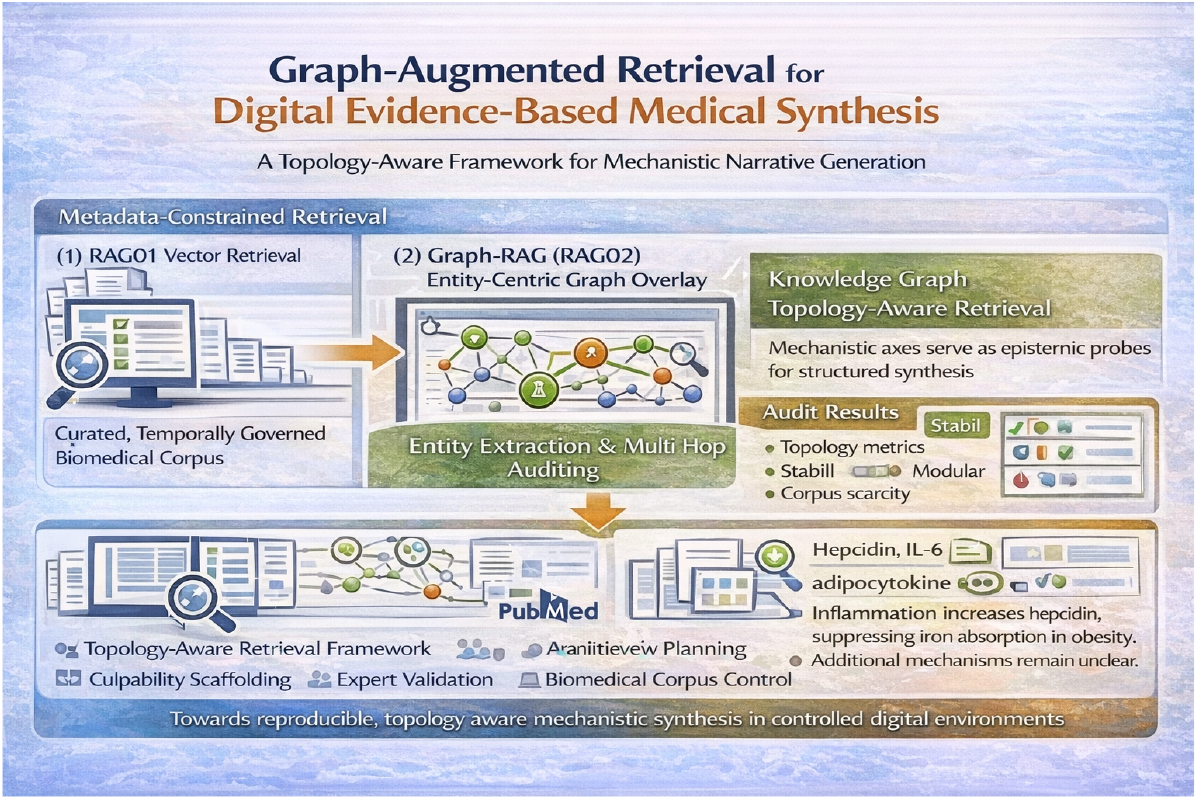
Experimental configuration and topology-aware evaluation workflow. Retrieval was performed under identical deterministic constraints (top-*k* = 5, cosine threshold = 0.50, publication year ≥ 2023) on a fixed corpus snapshot (11,861 chunks). RAG01 performs dense similarity ranking, while RAG02 integrates graph-aware scoring through weighted co-occurrence relationships derived from chunk-level entity extraction. Topology-aware diagnostics (local connectivity, induced subgraph density, modular overlap, and multi-hop stability) are used to classify mechanistic axes as structurally supported, weakly supported, or unsupported, distinguishing retrieval insufficiency from corpus-level evidentiary scarcity. The resulting configuration supports controlled mechanistic narrative synthesis within a reproducible environment.

#### Increased Iron Demand Axis

Demand-driven or volumetric mechanisms failed to demonstrate stable embedding at both 1-hop and 2-hop levels. Graph reinforcement was absent.

This axis was classified as unsupported within the indexed literature.

### 3.3 Local Topology and Modular Structure

Local topology was quantified on the core entity graph (30 nodes; 118 directed edges).

The 1-hop neighborhood size was larger for hepcidin than for obesity (16 vs 10 distinct neighbors), indicating broader local connectivity of the regulatory hub.

When evaluating induced 1-hop subgraphs (root node plus its neighborhood), obesity yielded an 11-node subgraph with 33 directed edges (density = 0.300), whereas hepcidin yielded a 17-node subgraph with 81 directed edges (density = 0.298). Thus, although hepcidin connects to a larger local neighborhood, edge density within these induced subgraphs remained comparable.

At 2-hop expansion, both nodes reached the majority of the graph (obesity: 27 nodes; hepcidin: 28 nodes), reflecting high global connectivity of the core topology.

The Jaccard overlap between obesity and hepcidin 1-hop neighborhoods was moderate (*J* = 0.444), indicating partial modular overlap while preserving distinct local neighborhoods.

Collectively, these metrics support a model in which obesity connects to the iron-regulatory core primarily through inflammatory mediators rather than through direct transporter or marrow-centered pathways.

### 3.4 Comparative Retrieval Behavior: RAG01 vs RAG02

Dense similarity retrieval (RAG01) identified semantically relevant material across all predefined mechanistic axes. However, similarity ranking alone did not reliably differentiate between structurally reinforced and weakly embedded mechanisms.

Across eight predefined mechanistic queries (top- *k* = 5; similarity threshold = 0.50; publication year ≥ 2023), graph-augmented retrieval (RAG02) preserved and slightly improved semantic alignment compared with RAG01.

Mean cosine similarity increased from 0.673 (SD 0.056) in RAG01 to 0.694 (SD 0.035) in RAG02. Graph activity ratio was 1.00 for both systems, indicating that under the contemporary filtered corpus snapshot, all retrieved top-*k* chunks were entity-annotated.

Importantly, graph-based scoring did not degrade semantic similarity. Rather, hybrid weighting introduced structural regularization, reducing similarity dispersion while preserving retrieval precision.

### 3.5 Integrated Mechanistic Synthesis

Across dense retrieval, graph-topological diagnostics, and modular analysis, only one pathway demonstrated consistent and stable structural embedding: inflammation-mediated hepcidin regulation.

Alternative mechanisms lacked reproducible reinforcement within the audited topology.

The objective of this analysis was not to identify novel biological relationships, but to evaluate whether corpus-derived topology faithfully reproduces established regulatory hierarchy and enables condition-level mechanistic discrimination.

The contribution therefore lies in methodological discrimination and topology-based validation rather than in the identification of previously unknown biological pathways.

## 4 Discussion

### 4.1 From Retrieval-Augmented Generation to Digital Evidence-Based Medicine

This study proposes a domain-constrained evolution of retrieval–augmented generation situated within a broader framework that may be described as digital evidence-based medicine (dEBM). While systems such as RAPID demonstrate that staged planning improves long-form generation, biomedical synthesis requires additional epistemic safeguards: corpus closure, temporal governance, mechanistic completeness, structural validation, and explicit discrimination between evidentiary absence and retrieval insufficiency.

In classical evidence-based medicine, validity derives from transparent integration of evidence with structured reasoning. Within a digital synthesis environment, these principles extend to computational governance of retrieval ranking, structural embedding, and narrative assembly.[15] The architecture presented here operationalizes these constraints through deterministic query planning, temporally validated retrieval, graph-theoretic auditing, and specialist review.

A key conceptual shift lies in redefining planning as mechanistic hypothesis interrogation.[9] Rather than merely expanding outline sections, topics are decomposed into explicit biological axes. Each axis functions as an epistemic probe whose classification (supported, weakly supported, unsupported) emerges from the interaction between dense similarity retrieval and structural embedding within a corpus-derived knowledge graph.

### 4.2 Topology-Aware Retrieval as Structural Validation

The Graph-RAG (RAG02) layer constitutes the principal methodological extension enabling topology-aware retrieval. In the obesity–iron deficiency case study, the audited entity network (30 nodes; 118 directed edges) displayed a centralized regulatory topology. Hepcidin occupied a high-connectivity position within the network, while obesity connected to the regulatory core primarily through inflammatory mediators such as IL-6. Multi-hop expansion consistently reproduced the sequence linking obesity, inflammatory signaling, hepcidin upregulation, ferroportin inhibition, and iron deficiency.[8]

Quantitatively, graph augmentation preserved semantic alignment and produced a consistent increase in mean cosine similarity while reducing similarity dispersion, indicating ranking stabilization rather than raw similarity inflation.

These findings indicate that hybrid scoring introduced structural regularization without degrading semantic relevance. Importantly, the underlying corpus and embedding space were unchanged, isolating topology-aware weighting as the differentiating factor [5].

Graph diagnostics enabled formal discrimination between narrative plausibility and structural reinforcement.[16] Hepcidin-independent transporter mechanisms, marrow remodeling pathways, and demand-driven hypotheses yielded semantically relevant excerpts under vector retrieval alone but failed to demonstrate stable 1-hop or 2-hop embedding within the knowledge graph. In this framework, absence of structural embedding reflects limited corpus-level reinforcement rather than retrieval failure. This distinction is foundational to dEBM: mechanisms lacking consistent topological support are explicitly classified as weakly supported or unsupported, rather than amplified through narrative extrapolation.

**Definition: Topology-Aware Retrieval in Biomedical Synthesis**

A retrieval framework is defined as *topology-aware* when ranking, mechanistic prioritization, and gap classification are influenced not only by semantic similarity between query and text, but also by the measurable structural embedding of concepts within a corpus-derived knowledge graph.

Topology refers to quantifiable relational properties of the indexed knowledge space, including node connectivity, weighted cooccurrence, induced subgraph density, modular overlap, and multi-hop stability. A topology-aware system therefore evaluates not only semantic relevance, but also structural reinforcement and integration within the corpus.

Structural prominence is not equated with biological truth. Rather, topology-awareness introduces a coherence constraint that enables transparent discrimination between semantically plausible mechanisms and those consistently reinforced across the literature topology.

### 4.3 Causal Scaffolding and Controlled Generation

Generation was further constrained by a biologically ordered causal scaffold linking condition, inflammatory signaling, regulatory nodes, iron flux, and phenotype. Concordance between this scaffold and the empirical graph topology suggests that the generative ordering reflects intrinsic structural organization of the indexed biomedical literature rather than arbitrary narrative imposition.

From a computational epistemology perspective, the architecture integrates four layers: similarity-based retrieval, graph-topological auditing, causal ordering, and specialist review. Together, these components approximate structured evidentiary reasoning within a controlled digital environment.

### 4.4 Closed Corpus Design and Reproducibility

The closed-corpus model enhances alignment with evidence-based standards by enforcing temporal governance, deterministic retrieval, and full provenance traceability, thereby mitigating risks analogous to retrieval-stage vulnerabilities documented in open RAG pipelines.[17] While open-domain retrieval maximizes breadth, it introduces instability and reproducibility challenges.[18] The present design prioritizes methodological transparency over exploratory dynamism, enabling audit-ready synthesis.

This closure may limit inclusion of emerging or niche evidence. However, explicit corpus boundaries and deterministic ranking support reproducible comparison between retrieval configurations and facilitate structural validation of mechanistic claims.

### 4.5 Limitations and Future Directions

Several limitations merit consideration. The entity network is derived from chunk-level co-occurrence rather than explicit causal extraction, and edge weights may partially reflect research intensity rather than biological magnitude. Structural absence does not exclude emerging plausibility, particularly for under-investigated pathways. The temporally filtered corpus (publication year ≥ 2023) may preferentially amplify contemporary regulatory narratives.

The present evaluation is intentionally focused on a single mechanistic domain to isolate structural behavior under controlled conditions. Broader multidomain validation remains a necessary next step.

Future extensions may incorporate relation-extraction models, bias normalization strategies, cross-condition comparative validation, and larger multi-topic query sets to assess generalizability of topology-aware retrieval beyond the present case study.

### 4.6 Implications for AI-Assisted Systematic Reviews

The framework has direct implications for AI-assisted systematic reviews. Traditional reviews rely on predefined search strategies and manual critical appraisal to ensure transparency and minimize bias.[19] Most current LLM-based approaches remain similarity-driven and lack explicit structural validation mechanisms.[20][12]

By integrating mechanistic axis decomposition with graph-topological auditing, the proposed architecture introduces a reproducible layer of structural validation. Mechanisms that fail to demonstrate stable embedding across multi-hop expansion can be explicitly classified as weakly supported within the indexed corpus. This approach does not replace formal systematic review methodology but may complement it by providing topology-aware interrogation of mechanistic hypotheses.

In this perspective, AI-assisted systematic review should not be framed as automated summarization, but as structured, topology-aware evidence interrogation under explicit corpus governance. Such an architecture advances retrieval-augmented generation from a productivity tool toward a controlled and reproducible model of digital biomedical reasoning.

## Data Availability

All data produced in the present work are contained in the manuscript

